# England’s Lockdown vs. Sweden’s Herd Immunity: A Comparison of the Daily New COVID-19 Cases and Related Deaths Using Comparative Interrupted Time Series Analysis

**DOI:** 10.1101/2020.08.13.20174706

**Authors:** Moaath K. Mustafa Ali, Yazan Samhouri, Marwa Sabha, Lynna Alnimer

**Affiliations:** Marlene and Stewart Greenebaum Comprehensive Cancer Center, University of Maryland Medical Center, Baltimore, Maryland; Department of Hematology and Medical Oncology, Allegheny Health Network, Pittsburgh, Pennsylvania; Division of Rheumatology and Clinical Immunology, Department of Medicine, University of Maryland Medical Center, Baltimore, Maryland; Department of Internal Medicine, Ascension Providence Hospital, Michigan State University, College of Human Medicine, Southfield, Michigan

**Keywords:** England, Sweden, comparative interrupted time series, COVID-19, daily cases, mortality

## Abstract

**Background:** There is a lack of empirical evidence that lockdowns decrease daily cases of COVID-19 and related mortality compared to herd immunity. England implemented a delayed lockdown on March 23, 2020, but Sweden did not. We aim to examine the effect of lockdown on daily COVID-19 cases and related deaths during the first 100 days post-lockdown.

**Methods:** We compared daily cases of COVID-19 infection and related mortality in England and Sweden before and after lockdown intervention using a comparative-interrupted time series analysis. The period included was from COVID-19 pandemic onset till June 30, 2020.

**Results:** The adjusted-rate of daily COVID-19 infections was eight cases/10,000,000 person higher in England than Sweden before lockdown order (95% CI: 2-14, P=0.01). On the day of intervention (lagged lockdown), England had 693 more COVID-19 cases/10,000,000 person compared to Sweden (95% CI: 467-920, P<0.001). Compared to the pre-intervention period, the adjusted daily confirmed cases rate decreased by 19 cases/ 10,000,000 person compared to Sweden (95% CI: 13-26, P<0.001). There was a rate excess of 1.5 daily deaths/ 10,000,000 person in England compared to Sweden pre-intervention (95% CI: 1-2, P<0.001). The increased mortality rate resulted in 50 excess deaths/ 10,000,000 person related to COVID-19 in England compared to Sweden on the day of lockdown (95% CI: 30-71, P<0.001). Post-intervention, the rate of daily deaths in England decreased by two deaths/ 10,000,000 person compared to Sweden (95% CI: 1-3, P<0.001). During phases one and two of lockdown lifting in England, there was no rebound increase in daily cases or deaths compared to Sweden.

**Conclusion:** The lockdown order implemented in England on March 23, 2020, effectively decreased the daily new cases rate and related mortality compared to Sweden. There was no short-term increase in COVID-19 cases and related-deaths after the phases one and two of the lifting of restrictions in England compared to Sweden. This study provides empirical, comparative evidence that lockdowns slow the spread of COVID-19 in communities compared to herd immunity.

## Introduction

On January 12, 2020, the World Health Organization (WHO) confirmed that a novel coronavirus strain is the cause of a respiratory illness in Wuhan, China. As of June 3, WHO reported 277,989 and 38,589 confirmed cases of coronavirus disease-2019 (COVID-19) and 39,369 and 4,468 deaths in the United Kingdom (UK) and Sweden, respectively [1]. The first two cases of COVID-19 in England were reported on January 31, while the first death was reported on March 2 [2, 3]. On March 23, the UK government announced a complete lockdown to halt the virus spread, which included limiting many of the daily activities like traveling and shopping.

On January 31, Sweden confirmed its first case of COVID-19. The first death was reported on March 11 [4]. Unlike the UK, Sweden has not imposed a lockdown. The Public Health Agency issued recommendations to work from home if possible, avoid unnecessary travel, engage in social distancing, and stay at home for people above 70 and for people who have even minimal symptoms that could be related to COVID-19. Sweden did not close its preschools or elementary schools as a preventive measure, which was highly criticized by the media [5]. Unlike the UK, the “recommendations” by the Swedish government were not enforced by law, since the Swedish constitution prohibits ministerial rule and mandates that the Public Health Agency must initiate all necessary actions to prevent the spread of COVID-19 [6].

England and Sweden went in two very different directions; a delayed but complete lockdown in England and a more relaxed approach without complete lockdown in Sweden. Many researchers assessed the impact of lockdown on daily COVID-19 cases using predictive models or measuring changes in reproduction number [7-12]. On the other hand, several newspapers warned that lockdowns were ineffective in halting COVID-19 spread [13-15]. Meunier reported that there was no evidence of discontinuity in COVID-19 epidemic growth rate or reproduction rate trends before and after implementing full lockdowns in Italy, France, Spain, and the UK [16]. Moreover, Meunier reports that countries, where less restrictive policies implemented, had similar trends in pandemic evolution to countries where lockdowns implemented [16].

Contradictory and potentially biased messages about lockdown efficacy may confuse the public, policymakers, and medical community about the potential efficacy of lockdowns in controlling the pandemic spread. There is a lack of studies providing comparative data on lockdown efficacy in halting COVID-19 spread. One of the methods to evaluate the causal effect of an intervention is to use a quasi-experimental study design called interrupted time series (ITS) analysis [17]. The outcome of interest is collected at many time points before and after an intervention. This design measures the effect of intervention by analyzing the trend in outcome over time using regression [17]. Interrupted time series analysis is one of the rigorous methods to estimate the effect of interventions when randomization is not possible or unethical [18]. This design is particularly useful when health policy is implemented or in natural experiments [18]. Herein, we study the effect of lockdown order in England on the daily number and rate of newly diagnosed cases and deaths secondary to COVID-19 in the first 100 days after lockdown order using an ITS analysis in comparison to Sweden.

## Methods

### Study design, setting, and participants

We conducted a comparative ITS to assess the causal effect of lockdown implemented in England on the number of new cases and deaths secondary to COVID-19 in comparison to Sweden. The study protocol was reviewed by the Alleghany Health Network Institutional Review Board. Because the study used de-identified and publicly available data, it was considered exempt category.

### Variables and Data source

We obtained dates and daily counts of new cases and deaths from publicly accessible datasets. The Public Health England agency publishes laboratory-confirmed newly diagnosed cases and deaths of COVID-19, which is updated daily [19]. The Department for Health and Social Care combines counts from the UK National Health Service mass testing and commercial partners testing [19]. Death counts included patients who tested positive with COVID-19 and died, regardless of the cause of death [19]. Death counts are the sum of deaths reported by National Health Service, Public Health England Health Protection teams, and by linking data from multiple sources as explained elsewhere [19]. In order to obtain rates adjusted for population size, we used England’s total population count in 2018. The Public Health Agency recommends using the total population count based on 2018 mid-year estimates from the Office for National Statistics [20].

The Public Health Agency of Sweden reports new cases and deaths secondary to COVID-19, which is updated daily [21]. The numbers reported are based on laboratory-confirmed cases under the Infectious Disease control act. Death counts included patients who tested positive with COVID-19 and died, regardless of the cause of death [21]. We adjusted the number of new cases and deaths using the Swedish population estimate in the third quarter of 2018 [22].

### Statistical analysis

Data used in the analysis included dates, daily counts of new cases and deaths as continuous variables, intervention as a dummy variable (before/after intervention), and control as a dummy variable (England/Sweden). We fit regression models using dates as the independent variable and daily counts and deaths as the dependent variables. For daily new cases time series, we analyzed confirmed cases counts between February 23, 2020, and June 30, 2020. For the death time series, we analyzed death counts between March 7, 2020, and June 30, 2020. For further details regarding the regression equation used for analysis, refer to online supplementary.

Both linear and nonlinear polynomial models were fit for time series. We selected models using performance metrics. Models with the highest R-squared and lowest Akaike’s Information Criteria were used. Generalized least square regression models provided by the “nlme” R software package allow for errors to correlate or have unequal variance [23]. We accounted for variance secondary to autocorrelation using autoregressive and moving average models. In these models, previous time points predict the current time point. We used the Durbin-Watson test to examine for autoregression [24]. Autocorrelation function and partial autocorrelation function plots allowed to examine the autocorrelation. We modeled time series using different orders of autoregression (p) and moving averages (q) and chose models with the lowest Akaike’s Information Criteria.

Because the expected change in outcome lags behind the lockdown due to the biologic behavior of the virus, we shifted the intervention time point in the daily cases model and mortality model [25, 26]. In the model of the daily cases, we shifted the time of intervention five days from March 23, 2020, which is the median incubation period of COVID-19 [26]. In the model of daily deaths, we shifted the time of intervention by eighteen days, which is the sum of the median incubation period of 5 days and the median time from symptom onset to death of 13 days [25, 26]. In this paper, we use lockdown and lagged lockdown interchangeably to indicate the time of intervention.

We investigated the trends in COVID-19 cases and related deaths during phase one and phase two of lockdown reversal in England compared to Sweden using a similar analytic approach as described above.

We calculated the thirty-day incidence rate of confirmed cases and deaths in both countries and compared them using the median unbiased estimation. We used the “EpiTools” R statistical package to calculate the incidence rate ratio [27]. The confirmed cases count included all patients who tested positive for a thirty-day duration, starting five days post lockdown. The death count included all patients who died in a thirty-day duration, starting eighteen days post lockdown. For each country, we obtained case fatality by dividing the sum of all deaths in thirty days, starting eighteen days post lockdown by the sum of all newly diagnosed cases for thirty days period starting five days post lockdown. Population size was adjusted to ten million for both countries in figures and tables for ease of interpretation and because Sweden’s population was estimated 10,207,086 in mid-2018.

Regression diagnostics were used to evaluate model assumptions. All statistical tests were two-sided. Tests with p-value <0 05 were considered statistically significant. Investigators used R statistical software (version 3.6.3) for analysis [28].

## Results

Figures 1 and 2 show daily confirmed COVID-19 cases and daily COVID-19 related deaths in England and Sweden adjusted to population size.

In the post-lockdown period, the thirty-day incidence rate of confirmed cases in England was 20,071 cases/10,000,000 person and 15,181 cases/10,000,000 person in Sweden, with a relative incidence rate of 1.32 (P<0.001). The thirty-day incidence rate of deaths in England was 3,444 deaths/10,000,000 person and 2,417 deaths/10,000,000 person in Sweden, with a relative incidence rate of 1.42 (P<0.001). Case fatality in England was 17% and in Sweden, 16%.

Figures 3 and 4 show ITS analysis to estimate the effect of lockdown on adjusted daily COVID-19 cases and COVID-19 related deaths in England compared to Sweden adjusted to population size. In the pre-lockdown period, England experienced a higher adjusted-rate of daily COVID-19 infections by eight cases/10,000,000 person than Sweden (95% CI: 2-14, P=0.01). On the day of intervention (lagged lockdown), England had 693 more COVID-19 cases/10,000,000 person compared to Sweden (95% CI: 467-920, P<0.001). Compared to the pre-intervention period, England had a lower adjusted-daily confirmed case rate by 19 cases/ 10,000,000 person compared to Sweden (95% CI: 13-26, P<0.001). In the mortality model, there was a rate excess of 1.5 daily deaths/ 10,000,000 person in England compared to Sweden pre-intervention (95% CI: 1-2, P<0.001). The increased mortality rate resulted in 50 excess deaths/ 10,000,000 person related to COVID-19 in England compared to Sweden on the day of lockdown (95% CI: 30-71, P<0.001). Post-intervention, the rate of daily deaths in England decreased by two deaths/ 10,000,000 person compared to Sweden (95% CI: 1-3, P<0.001). For further details regarding regression models, refer to eTables 1 and 2. eFigures 1 and 2 show the trend and relation of COVID-19 daily cases and related deaths with lockdown, and both phases 1 and 2 of lifting restrictions in England compared to Sweden. There was no increase in COVID-19 daily cases or deaths during the lifting restrictions phase one and two in England compared to Sweden (eTable 1 and eTable 2).

## Discussion

Governments follow different approaches when it comes to tackling viral pandemics. One controversial way is to allow for herd immunity, which is a form of indirect protection from infections that occurs when a large percentage of the population has become immune to infection through previous exposure, thereby protecting non-immune individuals [29]. Supporters of this opinion believe that lockdown does not prevent the spread of viruses; it only delays the inevitable spread at the cost of severe social and economic losses. On the other hand, many argue that lockdowns decrease the reproduction number, which is the average number of secondary cases each case generates, hence decreasing or eliminating human to human transmission, deaths, and burden on the health system [30].

At the beginning of the COVID-19 pandemic, England and Sweden followed the herd immunity approach to control the spread. Ferguson et al. at Imperial College London published a report estimating that 510,000 people would die in the UK without any mitigation [30]. The UK prime minister announced a complete lockdown on March 23, 2020, while Sweden continued with the herd immunity. Table 1 shows the difference between England regulations and Sweden recommendations during the COVID-19 pandemic [31, 32]. To understand the impact of the UK lockdown on COVID-19 spread, we used an ITS and compared England outcomes with Sweden as a control due to lack of lockdown. This design is useful to assess the effect of implemented health policy or in natural experiments [18]. Authors elected to compare only England to Sweden because its health care system is the only one that is accountable to the UK government. Wales, Scotland, and Northern Ireland have their own publicly funded health care systems that are accountable to their governments [33]. Moreover, there is some variation in the health system and policies between the four countries [34]. Although most European countries implemented lockdowns, authors compared the UK to Sweden because of the delayed lockdown in the UK, which led to the rapid spread of the virus in the UK, similar to Sweden in the pre-lockdown phase. The ‘similar’ trends in the pre-lockdown are essential to draw inferential estimates.

**Table 1.** Comparison of measurements implemented by England and Sweden government to decrease the spread of COVID-19 infection

|  | England[31] | Sweden[32] |
| --- | --- | --- |
| Stay at home order | -People should only leave or be away from their home for very limited purposes:<br>1. Shopping for basic necessities, for example, food and medicine, which must be as infrequent as possible.<br>2. One form of exercise a day, for example, a run, walk or cycle alone or with members of one's household.<br>3. Any medical need, including to donate blood, avoid injury or illness, escape the risk of harm, or to provide care or to help a vulnerable person.<br>4. Traveling for work purposes, but only where individuals cannot work from home. | -A ban on visiting all of the nation's care homes for older people has been in place since April 1, 2020.<br>-Maintain social distancing by keeping a distance from each other and refraining from non-essential travel within the country. |
| Closing certain businesses and venues | -The government closed the following businesses to the public by law:<br>1. Pubs, cinemas, and theatres.<br>2. Clothing and electronics stores; hair, beauty and nail salons, and outdoor and indoor markets (not selling food).<br>3. Libraries, community centers, and youth centers.<br>4. Indoor and outdoor leisure facilities such as bowling alleys, arcades, and soft play facilities.<br>5. Communal places within parks, such as playgrounds, sports courts, and outdoor gyms.<br>6. Places of worship (except for funerals).<br>7. Hotels, hostels, bed and breakfasts, campsites, caravan parks, and boarding houses for commercial/leisure use, excluding the use by those who live in them permanently and those who are unable to return home | - All venues must ensure that tables are spaced appropriately to avoid crowding.<br>- People must always be seated when consuming any food or beverages. |
| Stopping public gatherings | - Public gatherings of more than two people are prohibited. | - Ban public gatherings of >500 people, this was lowered to 50 people on March 29, 2020.<br>- Upper secondary school and University education are conducted online, but preschools and elementary schools remained open. |
| Going to work | - People are allowed to travel for work purposes, including to provide voluntary or charitable services, where work from home is impossible. | - No specific recommendations, people were encouraged to refrain from non-essential travel. |
| Enforcing the law | - The police and local authorities have the power to enforce the requirements set out in law if people do not comply with them. If people breach these regulations, the police may:<br>- Instruct people to go home, leave an area, or disperse.<br>- Instruct people to take steps to stop their children from breaking these rules if they have already done so.<br>- Take an individual home or arrest an individual, where they believe it is necessary. | - The only recommendation that was subject to the penalty was banning public gatherings of more than 50; otherwise, everything else was a non-enforceable recommendation |
| Use of personal protective equipment including masks | - No recommendations.<br>- Start mandatory masks on public transportation on June 15, 2020. | - No recommendations. |

The estimated causal effect is the result of net anti-COVID-19 measurements implemented by England but not Sweden (Table 1). During the pre-lockdown phase, the COVID-19 epidemic was spreading at a higher rate in England than Sweden. The enhanced spread resulted in higher COVID-19 related mortality. After lockdown implementation, the rate of daily COVID 19 cases decreased in England in comparison to Sweden. The decline in daily confirmed cases rate after lockdown compared to pre-lockdown is estimated to be 19 fewer cases/ 10,000,000 person. Because COVID-19 cases dropped in England, both countries’ curves crossed over during May 2020 (Figure 1). Moreover, the daily COVID-19 mortality rate decreased by two deaths/10,000,000 in England compared to Sweden.

To prevent a rebound increase in COVID-19 spread, the UK used a planned timetable to lift restrictions using three phases [35]. Phase one started on May 13. During phase one, more people were allowed to go to work if needed, and to drive to outdoor spaces irrespective of the distance. Also, outdoor exercise time was increased. Phase two started on June 1. Children were able to return to early years setting, and for Reception. Year one and year six were allowed back to school in a smaller size. Non-essential retail stores were allowed to open gradually. More local transport in urban areas was allowed to open. Sporting events can take place behind closed doors. People were allowed to expand their household groups to include one other household. This gradual reopening was always associated with reinforcement of social distancing, wearing face-covering, and frequent handwashing. In this study, the trends of daily COVID-19 cases and related deaths did not increase in England compared to Sweden with phases one and two of lifting restrictions (eTable 2). Authors believe that this cautious, gradual reopening, in addition to people’s compliance with regulations, explains the absence of a second surge so far. Phase three started on July 4. However, the authors did not estimate COVID-19 daily cases and deaths in this phase.

Interrupted time-series analyses are difficult to interpret when outcomes follow a nonlinear relation. Commonly, epidemics grow exponentially, slow down, and then decay exponentially [36, 37]. This “natural evolution” was not seen universally in the COVID-19 pandemic. The authors used a comparative model to control for the complex and dynamic growth of the COVID-19 pandemic. Another method to estimate a health policy effect is to use difference-in-difference method [38]. Authors used comparative interrupted time series analysis because it was unclear if both countries would follow a parallel trend in the future and because interrupted time series allows for more complex time trends using nonlinear models [38, 39]. In addition to that, comparative time series analysis can adjust for non-parallel trends between groups and allows for time-varying confounders [39].

Innate to non-randomized designs, our study has some limitations. England and Sweden are geographically separated, hence other confounders or co-interventions may have affected the outcomes. Time-varying confounders, including seasonality, can threaten the internal validity of ITS analysis. Authors believe this is unlikely due to several reasons. Both countries used similar tools to measure outcomes [19, 21]. Second, England and Sweden have a comparable life expectancy (80.8 vs. 81.1 years, respectively) suggestive of similar health quality [40]. Lastly, the COVID-19 pandemic started at the same time in both counties. This study only addresses short-term outcomes of lockdown order; the authors analyzed data of the first 100 days post-lockdown. Long-term trends in COVID-19 infection were not addressed and may increase after phase 3 of lockdown loosening in England.

In conclusion, the lockdown implemented in England did result in a statistically and epidemiologically significant reduction in the COVID-19 daily new cases rate compared to Sweden. The decline in daily cases also resulted in a drop in daily death rate related to COVID-19 in England. Lockdown in England has successfully halted COVID-19 transmission in the short-term. However, long term outcomes are unknown, and a new epidemic during winter is possible. Future research is needed to understand the long-term net outcome of the anti-COVID-19 policy implemented in both countries.

## Data Availability

Publicly available

## Contributors

Dr. Mustafa Ali had full access to all the data and analysis in the study and takes responsibility for the integrity of data and the accuracy of the data analysis.

Concept and design: Mustafa Ali, Samhouri

Acquisition, data analysis, and result interpretation: All authors.

Drafting of the manuscript: all authors.

Critical revision of the manuscript: all authors.

Statistical analysis: Mustafa Ali.

Administrative and technical support: Mustafa Ali, Samhouri.

Supervision: Mustafa Ali, Samhouri.

## Funding

This study was unfunded.

## Competing interests

All authors have no conflict of interest.

## Ethical approval

The study protocol was reviewed by the Alleghany Health Network Institutional Review Board. Because the study used de-identified and publicly available data, it was considered exempt category.

## Data Sharing

Data is available publicly. Statistical analysis code is available on request.

## Transparency statement

Mustafa Ali affirms that the manuscript is an honest, accurate, and transparent account of the study being reported; that no important aspects of the study have been omitted; and that any discrepancies from the study as planned have been explained.

## List of abbreviations

UK: United Kingdom
ITS: interrupted time series

## Figure legends

*Figure 1 Daily new cases of COVID-19 in England and Sweden adjusted for population size*

* Population size was adjusted to ten million for both countries in this figure.

*Figure 2 Daily deaths related to COVID-19 in England and Sweden adjusted for population size*

* Population size was adjusted to ten million for both countries in this figure.

*Figure 3 Comparative time series analysis to assess the impact of England’s lockdown order on daily new cases of COVID-19 compared to Sweden adjusted for population size*

* Population size was adjusted to ten million for both countries in this figure. NB: Dashed lines represent predicted daily cases in an interrupted time series model for each country

*Figure 4 Comparative time series analysis to assess the impact of England’s lockdown order on daily deaths related to COVID-19 compared to Sweden adjusted for population size*

* Population size was adjusted to ten million for both countries in this figure. NB: Dashed lines represent predicted daily cases in an interrupted time series model for each country

## Notes

### Competing Interest Statement

The authors have declared no competing interest.

### Funding Statement

None

### Author Declarations

Study is IRB exempt

